# Estimating changes in life expectancy in Hong Kong during the COVID-19 pandemic

**DOI:** 10.1101/2024.12.12.24318910

**Authors:** Alexandra H. T. Law, Anne M. Presanis, Justin K. Cheung, Peng Wu, C. Mary Schooling, Benjamin J. Cowling, Jessica Y. Wong

## Abstract

**Background:** Hong Kong has one of the longest life expectancies in the world but was heavily impacted by COVID-19 in 2022.

**Methods:** We constructed sex-specific life tables from 1998-2023 using parametric bootstrapping to account for statistical uncertainty in mortality rates. We used Arriaga’s decomposition method to estimate age-specific contributions to overall changes in life expectancy over this period. We also estimated cause-specific mortality rates.

**Results:** Hong Kong reported 50666 deaths in 2020, 51354 in 2021, 63692 in 2022, and 54731 in 2023. Estimates of life expectancy in males and females in 2020 and 2021 were similar to the pre-pandemic trend from 1998-2019 but declined significantly in 2022. Life expectancy for males was 82.3 years in 2021 and 80.4 years in 2022, and for females it was 87.9 in 2021 and 86.4 years in 2022. Compared to the pre-pandemic trend, the 2022 values corresponded to reductions by 2.22 (95% CI: 2.08, 2.36) years in males and 2.30 (95% CI: 2.17, 2.43) years in females. The loss in life expectancy in 2022 was mainly attributed to increased respiratory mortality rates in adults aged 65 or above. In 2023 life expectancy increased by 0.60 (95% CI: 0.46, 0.75) years in males and by 1.10 (95% CI: 0.95, 1.26) years in females.

**Conclusions:** In 2022 a very high respiratory mortality rate in older adults in Hong Kong during the COVID-19 pandemic was associated with a reduction in life expectancy by more than 2 years. In 2023 life expectancy increased towards the pre-pandemic trend.

## INTRODUCTION

Life expectancy from birth is a common measure used for international comparisons of population health ^1^. With advancement in healthcare resulting from successful economic development, Hong Kong’s life expectancy has risen significantly over the decades, making it one of the longest in the world ^2^. A decrease in life expectancy was observed in several Asian countries with high life expectancies in 2022 after the spread of Omicron, including Japan ^3^ and South Korea ^4^.

Hong Kong used public health and social measures to minimize COVID-19 transmission for the first two years of the pandemic ^5^, but experienced a major epidemic of Omicron BA.2 in early 2022 resulting in around half of the population being infected and more than 10,000 confirmed COVID-19 deaths among Hong Kong’s 7.3 million population ^6,7^. The majority of the deaths were observed in older adults ≥65 years of age ^8^. The reported number of COVID-19 deaths might not reveal the full picture of the impact of the pandemic because some confirmed deaths might have occurred with COVID-19 rather than because of COVID-19, while the control measures used to reduce transmission also had a substantial indirect impact which may have affected mortality ^9^. The World Health Organization estimated that 14.83 million excess deaths occurred in the first two years of the pandemic ^10^. The Institute for Health Metrics and Evaluation estimated that global life expectancy decreased by 1.6 years from 2019 to 2021 ^11^, which was a dramatic reversal from the slowly increasing trend before 2019.

To explore how the COVID-19 pandemic affected mortality in Hong Kong, a city having a high proportion of older adults, in this study we estimated patterns in mortality rates and the consequent life expectancy changes in Hong Kong during the COVID-19 pandemic.

## METHODS

### Sources of data

Age-specific weekly deaths from 1998 to 2023 were obtained from the Census and Statistics Department of the Government of the Hong Kong Special Administrative Region. For analysis of cause-specific death rates, we selected 9 major primary causes of death, including malignant neoplasms, cardiovascular, respiratory, kidney diseases, diabetes mellitus, chronic liver disease, dementia, septicemia, and external causes. A final category, “other causes”, included all other deaths. These ten cause of death groupings were coded according to the International Classification of Diseases, Ninth Revision (ICD-9) and Tenth Revision (ICD-10) (Appendix). Almost all deaths in Hong Kong occur in hospital, which facilitates accurate coding. The age-specific mid-year population sizes from 1998 to 2023 were obtained from the Census and Statistics Department and were used as the denominators for the estimation of mortality rates ^12^. The population sizes for ages 85 to ≥100 years in 1996, 2001, 2006, 2011, 2016, and 2021 were obtained from the censuses and by-censuses conducted by the Census and Statistics Department ^13^. These data were used to estimate the population size from age 85 to ≥100 years from 1998 to 2023 (Appendix). Our study received ethical approval from the Institutional Review Board of the University of Hong Kong.

### Statistical analysis

We constructed life tables for each year from 1998 to 2023 based on local mortality rates by age and sex (Appendix). We used a parametric bootstrap approach assuming that age- and sex-specific mortality rates followed Poisson distributions, allowing us to construct uncertainty intervals for the estimated life expectancy. We used simple linear regression models to fit the change in life expectancy between 1998 and 2019 and extrapolated this linear fit to the pandemic years.

Arriaga’s decomposition method was used to investigate the contribution of different age groups to changes in life expectancy in 2020, 2021, and 2022 ^14^. The age-specific contribution consists of both direct and indirect effects taking the preceding year as a reference. However, the last age group only gives a direct contribution, as there is no older age constituting an indirect effect. The sum of effects from all ages and the total change in life expectancy in years should be equal. The change in life expectancy by age was estimated for six age groups: 0-4, 5-14, 15-44, 45-64, 65-79, and ≥80 years. We did not conduct this decomposition analysis for 2023 versus 2022 because the very high mortality rates in 2022 would affect the comparison. All statistical analyses were conducted in R version 4.3.3 (R Foundation for Statistical Computing, Vienna, Austria).

## RESULTS

Figure 1 shows the all-cause mortality rates from 1998 to 2023 for males and females in six age groups. From 1998 through to 2021 the age-specific mortality rates generally declined over time for both males and females. The mortality rates increased substantially in 2022 in all age groups except in 5-14 years. In particular, the estimated mortality rates among males and females ≥80 years increased dramatically from 2021 to 2022. For males, mortality rates increased from 8120 per 100,000 in 2021 to 10800 per 100,000 in 2022, and for females it increased from 6590 per 100,000 in 2021 to 8340 per 100,000 in 2022. The long-term decreases in mortality rates over the study period resulted in a steady increase in the life expectancy of both sexes, from 77.3 in 1998 to 82.3 in 2021 for males, and from 83.1 in 1998 to 87.9 in 2021 for females in Figure 2. In 2022, life expectancy declined to 80.4 years in males and 86.4 years in females. Using linear regression models, we projected the trend in life expectancy from 1998 through 2019 into the three pandemic years, shown as dotted lines in Figure 2. Compared to those secular trends in males and females, life expectancy decreased slightly in 2020 and 2021 in females while it remained similar to the projected trend in males in those two years. Life expectancy then declined very substantially in 2022 by 2.22 (95% confidence interval, CI: 2.08, 2.36) years in males and by 2.30 (95% CI: 2.17, 2.43) years in females, compared to the extrapolated value from the slowly increasing trend from 1998-2019.

**Figure 1:**
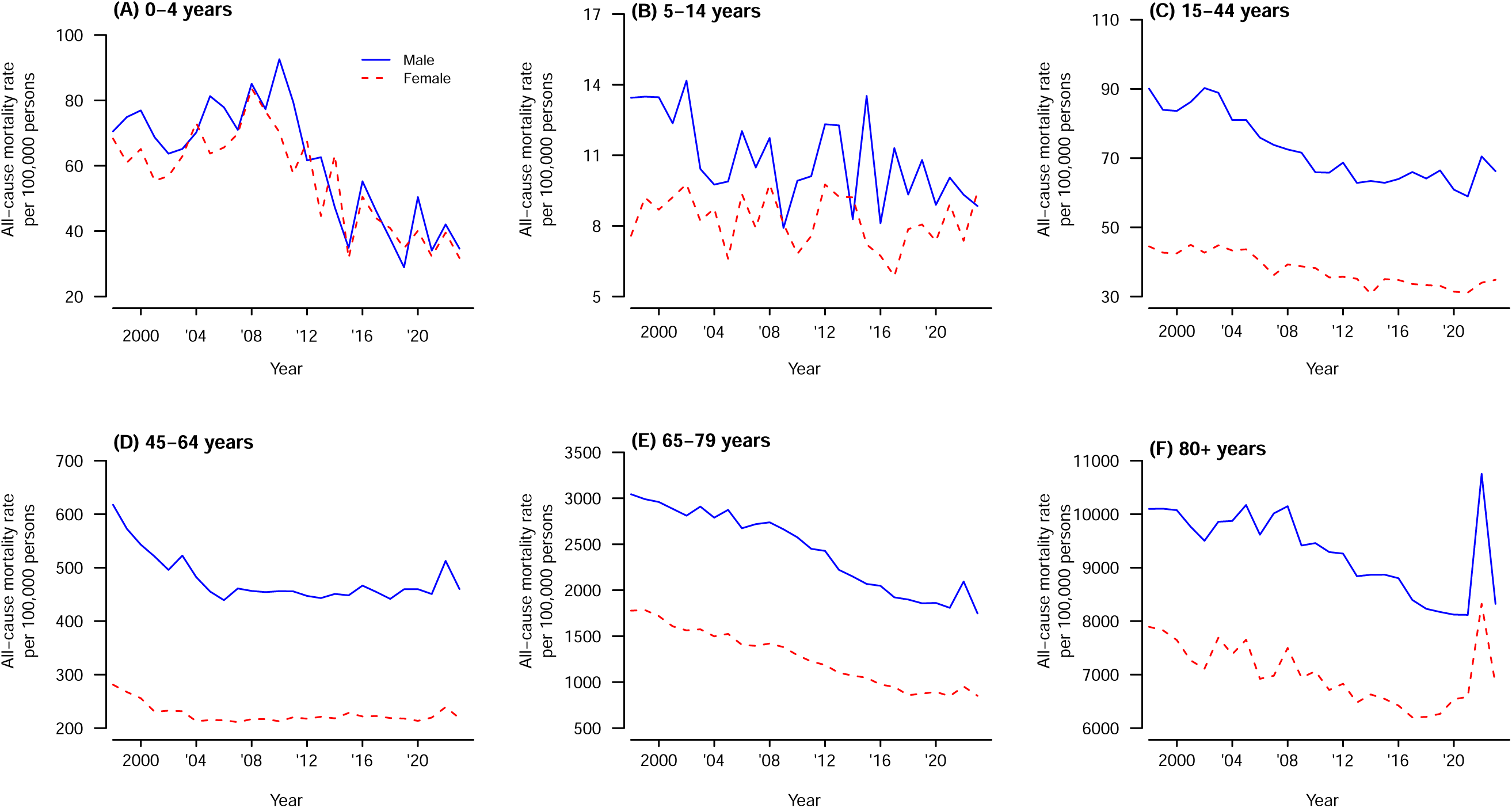
All-cause mortality rates in Hong Kong by age and sex, from 1998-2023.

**Figure 2:**
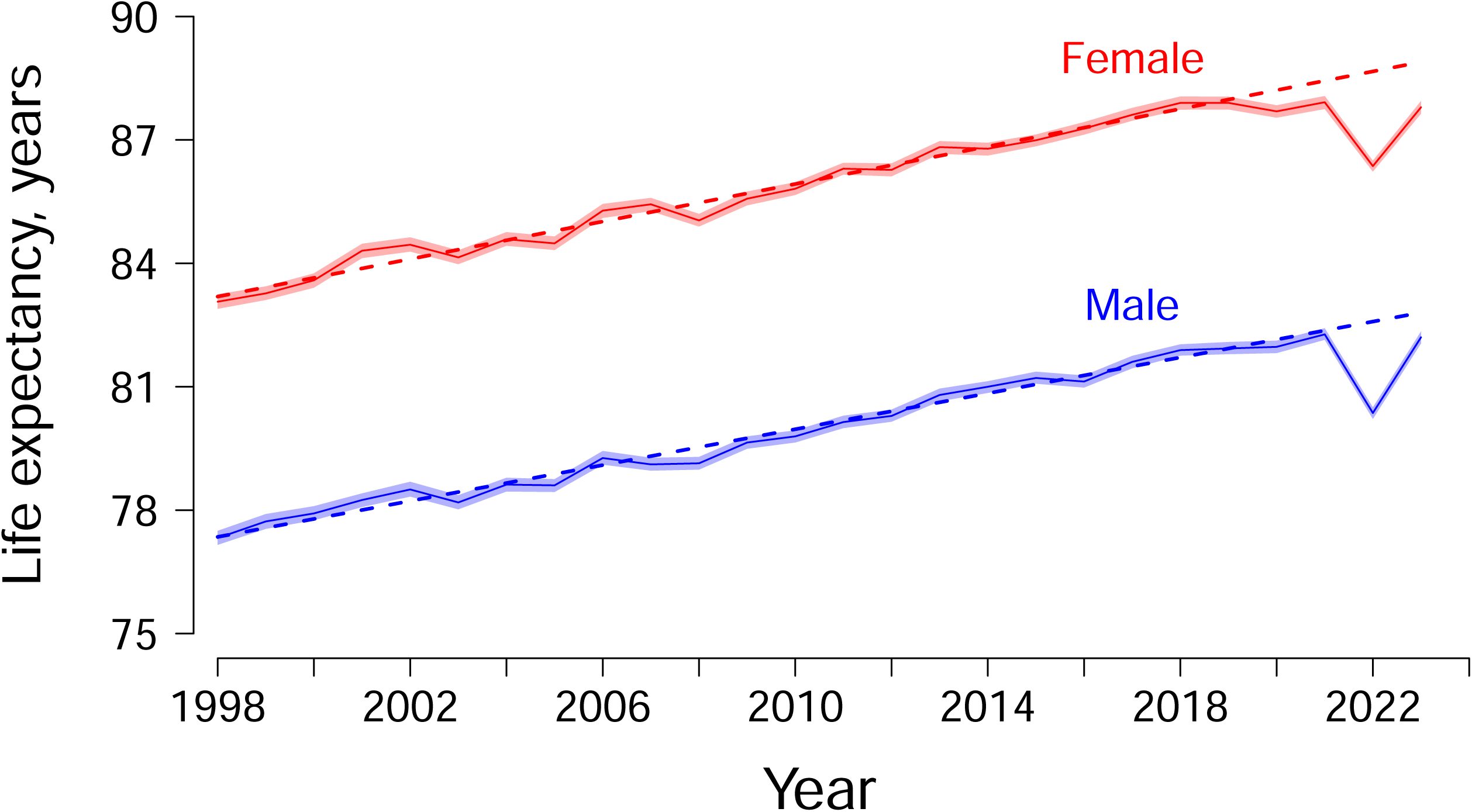
Life Expectancy for males and females in Hong Kong from 1998-2023 with 95% confidence intervals. The dashed lines show the fitted linear trends from 1998-2019 extrapolated to 2020-23.

In 2023, life expectancy rose in males and females back towards the pre-pandemic trend, increasing by 0.60 (95% CI: 0.46, 0.75) years in males and by 1.10 (95% CI: 0.95, 1.26) years in females. The life expectancy estimates of 82.2 (95% CI: 82.1, 82.3 years in males and 87.8 (95% CI: 87.6, 87.9) years in females were still lower than the pre-pandemic trend which under our simple linear extrapolation would have corresponded to life expectancies of 82.8 years in males and 88.9 years in females in 2023 (Figure 2).

In our decomposition analysis by age group, the statistically significant reduction in female life expectancy from 2021 to 2022 was largely attributed to the increase in mortality rate among females aged 80 years or above, followed by those aged 65-79 years (Figure 3). Similarly, the decrease in male life expectancy from 2021 to 2022 was also attributed to increased mortality rates in males aged ≥80 years, followed by those 65-79 years.

**Figure 3:**
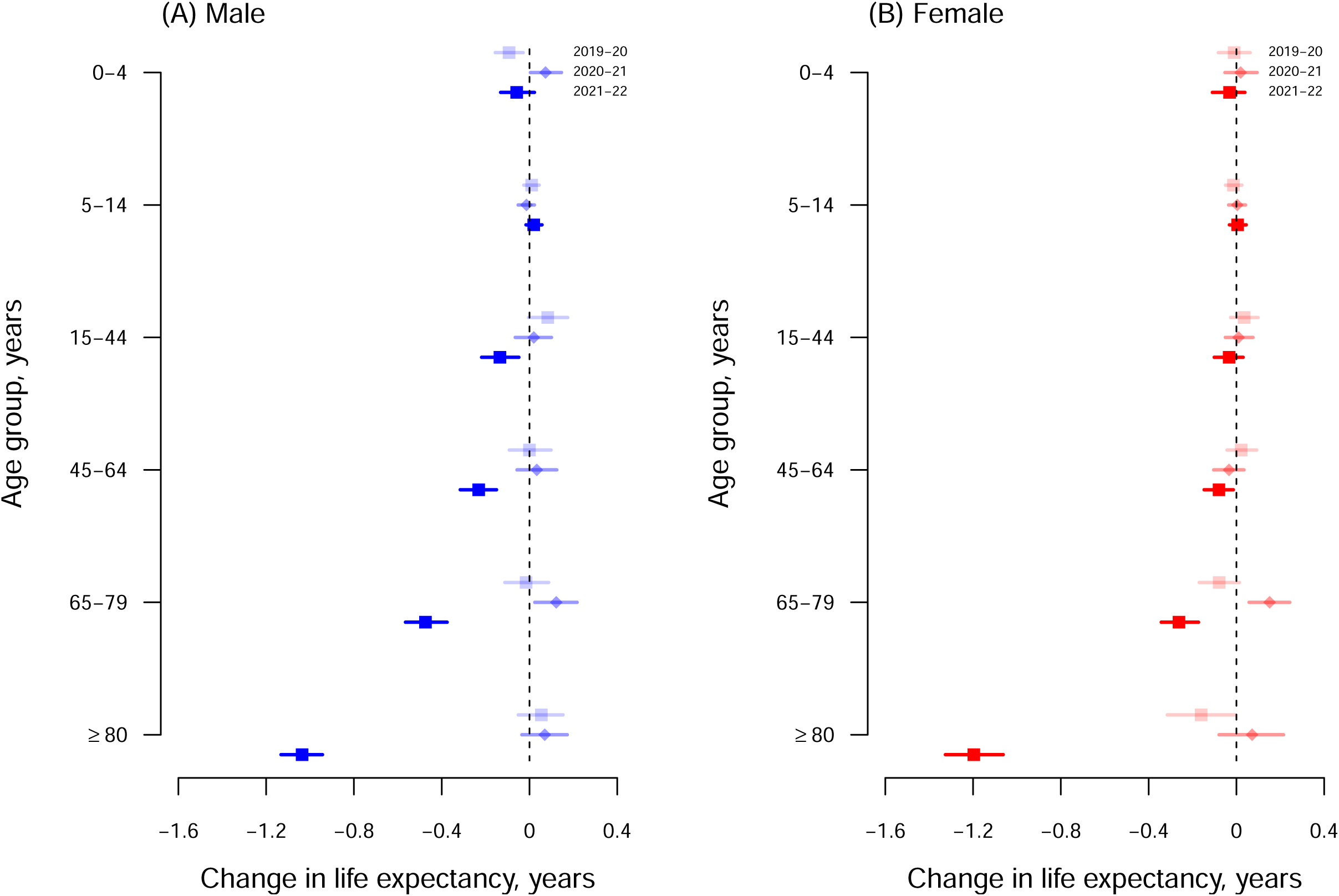
Arriaga decomposition of age-specific contributions (with 95% confidence intervals) to changes in life expectancy in 2019-2020 (light-shaded squares), 2020-2021 (diamonds), and 2021-2022 (dark-shaded squares) for males and females. Each estimate shows the positive or negative change in life expectancy in the corresponding age group between consecutive years.

Figure 4 shows the mortality rates by cause and by sex from 1998-2023. The largest increase in 2022 was observed in mortality from respiratory causes, and smaller increases in mortality rates were observed in 2022 in several other cause of death categories, except cancer for males, in 2022. From 1998-2023, most of the cause-specific mortality rate of males were higher than that of females, apart from diabetes, kidney diseases, and septicemia. In 2023, cause-specific mortality rates returned to levels that were comparable to those prior to the COVID-19 pandemic for some causes although respiratory and cardiovascular mortality remained elevated.

**Figure 4:**
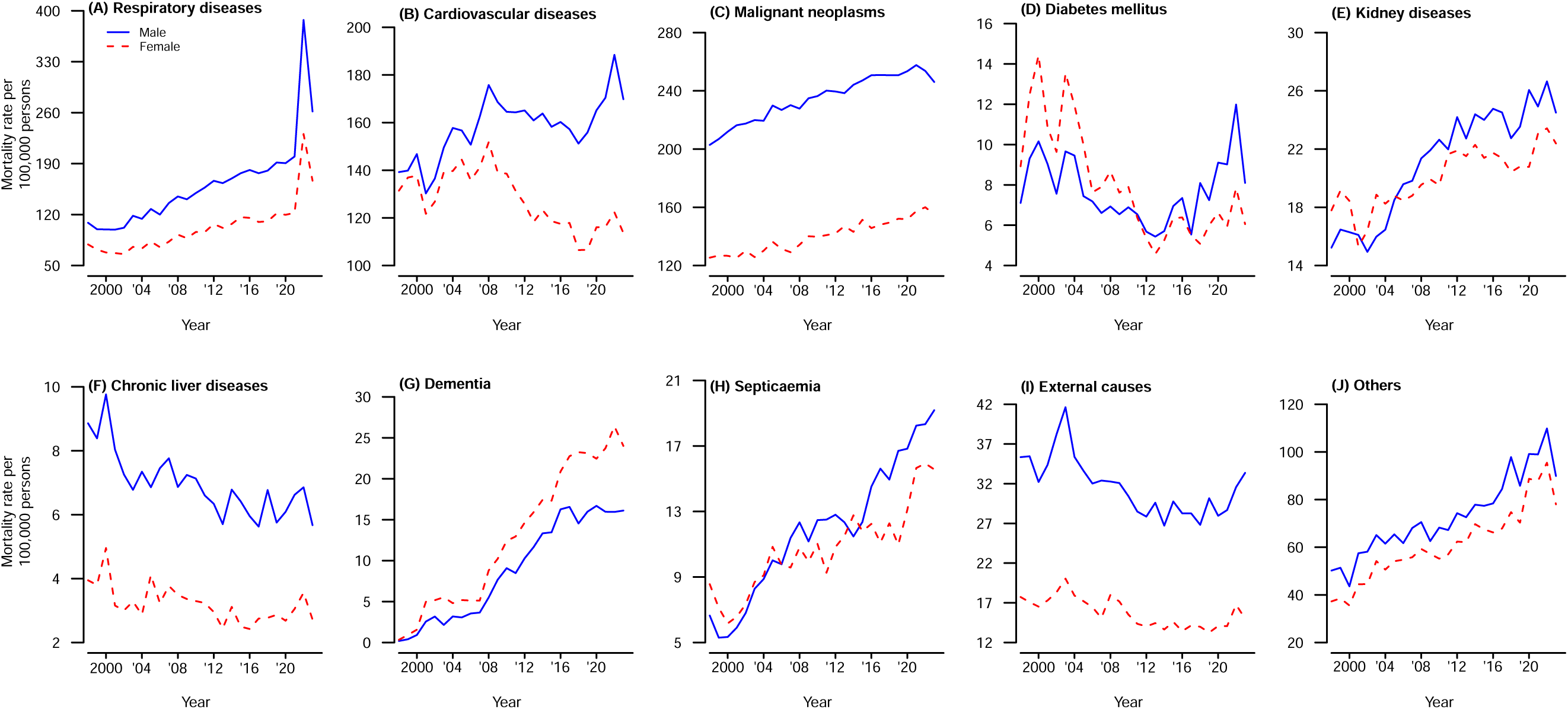
Mortality rates in Hong Kong by cause and by sex from 1998-2023.

## DISCUSSION

COVID-19 was successfully contained in Hong Kong in 2020 and 2021, with fewer than 1% of the population infected ^15^, but the uncontrolled spread of Omicron BA.2 in early 2022 had a very substantial impact ^6,7^. The loss in life expectancy in 2022 compared to an extrapolated prediction based on the pre-pandemic trend, by 2.22 years in males and by 2.30 years in females, was mainly attributed to a large increase in mortality rates in persons aged ≥65 years (Figure 3). In cause-specific analysis we identified increases in mortality from respiratory causes and cardiovascular causes (Figure 4). There was no evidence of substantial displacement of mortality from other causes. In 2023, life expectancy in Hong Kong increased towards the pre-pandemic trend (Figure 2), but mortality rates remained elevated for respiratory diseases and cardiovascular diseases (Figure 4) and male and female life expectancy remained somewhat below the extrapolated pre-pandemic trend.

The high mortality impact of SARS-CoV-2 Omicron BA.2 in Hong Kong in early 2022 has been discussed at length elsewhere, and can be attributed to lower COVID-19 vaccine coverage in older adults and strain on healthcare resources ^6,16,17^. By early 2022 the vaccination rate among individuals aged 80 years or above was the lowest among all adult age groups in Hong Kong, with 20.5% of males and 17.1% of females in this age group receiving at least two doses of SARS-CoV-2 vaccination at the beginning of 2022, compared to over 80% for adults ≥18 years overall ^5,18,19^. SARS-CoV-2 Omicron BA.2 infections spread explosively in early 2022, causing severe strain on healthcare resources leading to a three-fold increase in case fatality rates at the peak of the epidemic in March compared to earlier or later in that epidemic ^16^. The loss of slightly more than two years of life expectancy in both males and females during the COVID-19 pandemic in Hong Kong implies there were a substantial number of years of life lost.

Life expectancy is calculated from a life table where the observed age-specific mortality rates are applied to a standard population of one million births, as if the age-specific mortality rates would apply to that cohort as they progress through life. This allows a somewhat standardized comparison of age-specific mortality rates among population with different age structures. In Hong Kong’s ageing population the COVID-19 pandemic caused a very substantial number of deaths in older adults, and the COVID-19 mortality rate in Hong Kong in 2022 was one of the highest per capita rates in the world. Estimating the change in life expectancy enables a form of age-adjusted comparison with mortality rates in other locations.

Prior to the COVID-19 pandemic, global life expectancy increased from 62 years in 1980 to 72 years in 2015 ^11^. Hong Kong has also experienced steady increases in life expectancy over the last 25 years, but the COVID-19 pandemic had a drastic impact on life expectancy in 2022 (Figure 2). COVID-19 likely impacted life expectancy in almost every country, for example the life expectancy of males (females) in the United States decreased significantly by 2.1 (1.5) years in 2020, by a further 0.7 (0.6) years in 2021, and then increased by 1.3 (0.9) years in 2022 to a level still 1.5 (1.2) years below that in 2019 ^20^. The United Kingdom reported reductions of life expectancy by 0.1-0.3 years each consecutive year in 2020, 2021 and 2022 ^21^.

Locations with high COVID-19 mortality rates in 2020 and 2021 would tend to have reductions in estimated life expectancy in those years. In contrast, Hong Kong along with several other locations in the Asia-Pacific region was able to minimize COVID-19 mortality until 2022 ^5^ and did not have substantial changes in life expectancy until 2022. Similar to Hong Kong, South Korea had slight increases in life expectancy in 2020 and 2021 but then had a reduction in life expectancy by 0.7 years in males and 1.0 years in females in 2022 ^22^. Singapore reported almost no change in life expectancy in 2020, reductions by 0.4 and 0.5 years in males and females in 2021, and by 0.3 and 0.1 years in 2022 respectively ^23^. Australia and New Zealand reported almost no change in life expectancy over the period 2020-22 ^24,25^.

There are several limitations to discuss. First, we were not able to obtain mid-year population size for ages 85 to ≥100 years, and we made reasonable assumptions about the distribution of population in this range (Appendix). Second, Arriaga’s decomposition method is a common approach to assess changes in life expectancy but it is limited to decomposing the discrete changes such as changes from one year to the next. In Arriaga’s decomposition method, the timing of changes within the time intervals is not captured, and the effects are averaged over the discrete periods. Third, we were not able to separate the direct impact, referring to deaths from COVID-19, and indirect impact, referring to the consequences of intervention policies implemented during the pandemic for example delays in ambulances responding to heart attacks, or increases in underlying medical conditions due to social isolation, change of lifestyles and delay of healthcare seeking behaviors, etc. We previously reported an increase in cardiovascular mortality in 2020 likely attributable to changes in healthcare seeking behaviors ^9^. In a future study it would be of interest to separate the direct and indirect effects of the pandemic by including more covariates, including the effectiveness of the intervention policies, the long-term health impacts resulting from SARS-CoV-2 infections, and changes in access to and utilization of medical services.

In conclusion, we estimated a substantial decrease in life expectancy in males and females in 2022, associated with substantial increases in all-cause mortality rates particularly in males and females aged 80 years or above, followed by those aged 65 to 79 years, when a substantial drop in life expectancy occurred compared to the gradually increasing trend in the pre-pandemic years. Life expectancy increased somewhat in 2023 although it did not return to the pre-pandemic trend.

## Data Availability

Age-specific weekly deaths from 1998 to 2023 were obtained from the Census and Statistics Department of the Government of the Hong Kong Special Administrative Region. The age-specific mid-year population sizes from 1998 to 2023 were obtained from the Census and Statistics Department.

## ACKNOWLEDGEMENTS

The authors thank Julie Au for technical support.

