## Appendix for "Estimating changes in life expectancy in Hong Kong during the COVID-19 pandemic"

This appendix provides additional information on the data and statistical methods used within the study.

Table S1. ICD-9 and ICD-10 codes for underlying causes of death that were included in the analysis

| **Cause of death** | **ICD-9** | **ICD-10** |
| --- | --- | --- |
| Respiratory diseases | 460-519 | J00-J99 |
| Cardiovascular diseases | 390-459 | I00-I99 |
| Malignant neoplasms | 140-208 | C00-C97 |
| Diabetes mellitus | 250 | E10-E14 |
| Kidney diseases | 580-589 | N00-N07, N17-N19, N25-N27 |
| Chronic liver diseases | 571 | K70, K73-K74 |
| Dementia | 290 | F01-F03 |
| Septicaemia | 038 | A40-A41 |
| External causes | 800-999 | S00-T99 |

**Supplementary Methods**

1. **Distribution of old population aged 85 to 100 or above**

The population sizes for individuals of ages 85 to ≥100 years of age in 1996, 2001, 2006, 2011, 2016, and 2021 were obtained from the censuses and by-censuses conducted by the Census and Statistics Department ^1^. The population size from age 85 to ≥100 years in the non-census years from 1998 to 2021 were interpolated based on the number of people in each year of age from age 85 to ≥100 years between the above census years, while the population size in each year of age in 2022 and 2023 was estimated with the proportion of old adults in each year of age in 2021 multiplied by the total number of individuals aged ≥85 in 2022 and 2023 estimated by the Census and Statistics Department (Tables S2 and S3).

Table S2. Census data of male in 1996, 2001, 2006, 2007, 2011, 2016, and 2021 census. Population sizes for males of each year of age in non-census years 1997 through 2020 were estimated by interpolation from the two closest values in the table below, while estimates for 2022 and 2023 were obtained by multiplying the proportions in 2021 by the total number of ≥85y in 2022 and 2023.

|  | 1996 | 2001 | 2006 | 2011 | 2016 | 2021 | |
| --- | --- | --- | --- | --- | --- | --- | --- |
| Age, years | Number | Number | Number | Number | Number | Number | Proportion |
| 85 | 3337 | 3952 | 6277 | 7736 | 11464 | 12650 | 15% |
| 86 | 2063 | 3166 | 4877 | 6623 | 9717 | 11303 | 13% |
| 87 | 1638 | 2632 | 4068 | 5191 | 7365 | 10266 | 12% |
| 88 | 1036 | 2139 | 2758 | 4736 | 6503 | 9737 | 12% |
| 89 | 1194 | 1817 | 2803 | 3662 | 5301 | 9010 | 11% |
| 90 | 767 | 1514 | 1917 | 3063 | 4143 | 7030 | 8.3% |
| 91 | 434 | 925 | 1681 | 2549 | 3324 | 5555 | 6.6% |
| 92 | 397 | 760 | 1174 | 1851 | 2407 | 3902 | 4.6% |
| 93 | 339 | 533 | 815 | 1362 | 2306 | 3342 | 3.9% |
| 94 | 170 | 477 | 855 | 1188 | 1665 | 2594 | 3.1% |
| 95 | 146 | 347 | 488 | 792 | 1346 | 1867 | 2.2% |
| 96 | 135 | 275 | 361 | 608 | 1179 | 1495 | 1.8% |
| 97 | 70 | 217 | 144 | 263 | 748 | 943 | 1.1% |
| 98 | 24 | 170 | 126 | 300 | 503 | 1257 | 1.5% |
| 99 | 14 | 161 | 99 | 258 | 508 | 801 | 0.95% |
| 100+ | 56 | 125 | 358 | 519 | 534 | 2733 | 3.2% |

Table S3. Census data of female in 1996, 2001, 2006, 2007, 2011, 2016, and 2021 census. Population sizes for females of each year of age in non-census years 1997 through 2020 were estimated by interpolation from the two closest values in the table below, while estimates for 2022 and 2023 were obtained by multiplying the proportions in 2021 by the total number of ≥85y in 2022 and 2023.

|  | 1996 | 2001 | 2006 | 2011 | 2016 | 2021 | |
| --- | --- | --- | --- | --- | --- | --- | --- |
| Age, years | Number | Number | Number | Number | Number | Number | Proportion |
| 85 | 6341 | 7265 | 10620 | 12676 | 16233 | 15672 | 11% |
| 86 | 4489 | 6088 | 8441 | 11343 | 13932 | 15040 | 11% |
| 87 | 3353 | 5066 | 7666 | 10289 | 12517 | 13990 | 10% |
| 88 | 2773 | 4625 | 6594 | 9019 | 11583 | 13532 | 9.5% |
| 89 | 2786 | 4251 | 5526 | 7727 | 10542 | 12938 | 9.1% |
| 90 | 2370 | 3348 | 5587 | 7025 | 8713 | 11764 | 8.3% |
| 91 | 1609 | 2541 | 3994 | 5627 | 7588 | 9309 | 6.6% |
| 92 | 1424 | 2069 | 2928 | 4577 | 6693 | 8221 | 5.8% |
| 93 | 1240 | 1681 | 2703 | 3718 | 5642 | 7287 | 5.1% |
| 94 | 917 | 1341 | 2471 | 3400 | 4641 | 6437 | 4.5% |
| 95 | 763 | 1153 | 1740 | 2470 | 4168 | 4970 | 3.5% |
| 96 | 440 | 874 | 1220 | 1996 | 3022 | 4307 | 3.0% |
| 97 | 236 | 661 | 864 | 1428 | 2350 | 3707 | 2.6% |
| 98 | 237 | 498 | 653 | 1167 | 1782 | 3188 | 2.2% |
| 99 | 165 | 484 | 402 | 805 | 1751 | 2521 | 1.8% |
| 100+ | 389 | 572 | 1152 | 1371 | 3111 | 8842 | 6.2% |

**2. Construction of life tables using a parametric bootstrap**

Recognizing that there is uncertainty in mortality rates particularly in older age groups where the population sizes are smaller, we used a parametric bootstrap approach with a Poisson distribution. We generated 1000 sets of $q_{x,t}$, which were simulated using random draws from a Poisson distribution with the corresponding mortality rate and population size. These were then used to create 1000 life tables for each calendar year between 1998 and 2022. Our life tables ended at the open-ended age interval of 100 years ($L_{100+}$) for both males and females.

The standard life table parameters for each age x were estimated using the following equations:

$$l_{x}= l_{x-1} \times(1- q_{x-1})$$

$$d_{x}= l_{x} \times q_{x}$$

$$L_{x}= w_{x}l_{x+1}+ {a_{x}d}_{x}$$

$$T_{x}= \sum_{i=x}^{100+} L\left( x \right)$$

$$e_{x}= \frac{T_{x}}{l_{x}}$$

where$l_{x}$ is the number of survivors from birth to each age group x, $l_{x-1}$ is the number of survivors from birth to age group x-1, $q_{x-1}$ is the probability of death in age group x-1, $d_{x}$ is the number of deaths in age group x, $q_{x}$ is the probability of death in age group x, $L_{x}$ is the sum of person-years spent in age group x, $a_{x}$ is the average length of time to death for individuals who died in age group x, $w_{x}$ is the length of age group interval x, $T_{x}$ is the remaining number of person-years to live in age group x, and $e_{x}$ is the life expectancy in age group x. In particular, $e_{0}$ is the life expectancy at birth which is the outcome measure in our analyses.

In our analysis by individual age x, values of 1 and 0.5 were assumed for $w_{x}$ and $a_{x}$ respectively. In the first age group (age 0-1 years) we used $a_{x}$ of 0.068 as suggested by Preston et al. ^2^.

For the open-ended age interval, $L_{100+}$was estimated for males and females respectively using the following equations:

$$L_{100+,male}= exp(\gamma_{100+, male}+ \delta_{100+, male}\times d_{100+, male})$$

$$L_{100+,female}= exp(\gamma_{100+, female}+ \delta_{100+, female}\times d_{100+, female})$$

where $\gamma_{100+,male}$ and $\gamma_{100+, female}$ are the average number of person-years lived in age group 100+ for males and females$, \delta_{100+,male}$and $d_{100+, male}$ reflect changes in mortality in age group 100+ for males, and $\delta_{100+,female}$and $d_{100+, female}$ reflect changes in mortality in age group 100+ for females.

**3. Decomposition of changes in life expectancy**

The Arriaga decomposition method was used to investigate the contribution of different age groups to the life expectancy gap from 2020 to 2022 in Hong Kong ^3,4^. Total contribution of age group x to the change in life expectancy in year t+1 relative to year t ($C_{x})$can be expressed using the following equation:

$$C_{x=}\left[ \frac{l_{x}^{t}}{l_{0}}\left( \frac{L_{x}^{t+1}}{l_{x}^{t+1}}- \frac{L_{x}^{t}}{l_{x}^{t}} \right) \right]+ \left[ \frac{T_{x+1}^{t+1}}{l_{0}}\left( \frac{l_{x}^{t}}{l_{x}^{t+1}}- \frac{l_{x+1}^{t}}{l_{x+1}^{t+1}} \right) \right]$$

where $L_{x}$ is the number of person-years lived in age group x,$l_{0}$ is the number of survivors at age 0,$T_{x}$ is the total person-years lived at and beyond age group x, and $l_{x}$is the number of survivors from birth to age group x. The sum of the total contribution across all age groups should equal the total change in life expectancy from year t to year t+1.
